## Supplementary material for "Hospital-Level Variation in Antenatal Corticosteroids for Late Preterm Births": Online Supplement

**Supplemental Online Content**

**eMethods**

Identification of Maternal Delivery Hospitalizations

We identified maternal delivery hospitalizations using the Medicare Severity-Diagnosis Related Group Major Diagnostic Category classification system. We selected all hospital encounters classified as obstetric admissions related to pregnancy and childbirth. From these encounters, we extracted hospital identifier, admission date, and discharge date.

Gestational Age Classification

We extracted gestational age information for each maternal delivery encounter, using *International Classification of Diseases, Tenth Revision* (ICD-10) diagnosis codes. When multiple gestational age codes were present for a single encounter, we selected the maximum gestational age recorded during inpatient stay.

We classified deliveries into three gestational age categories:

- Early preterm: 24 to 33 weeks
- Late preterm: 34 to 36 weeks
- Term: 37 to 42 weeks

Delivery Date Determination

We implemented a hierarchical approach to establish a precise delivery date for each encounter. First, we identified *International Classification of Diseases, Tenth Revision, Procedure Coding System* (ICD-10-PCS) codes indicating vaginal or cesarean delivery procedures, selecting the latest procedure date within the admission window. Second, if procedure codes were unavailable, we used Current Procedural Terminology (CPT) codes for vaginal or cesarean delivery (see eTable 1). When neither source contained delivery codes, we used the discharge date as a proxy for delivery date. All procedure dates required to fall within the admission-to-discharge window; procedures recorded outside this window were excluded.

Antenatal Corticosteroid Exposure Identification

We identified antenatal late preterm steroid exposure using itemized billing records from the Premier charge master system, which contains detailed medication administration data. We matched billing records against a standardized list of betamethasone formulations designated for antenatal administration. To capture steroid administrations that occurred before or during the delivery hospitalizations – including cases when pregnancy spans calendar years – we incorporated billing files from both the index year and the preceding year.

For each patient with a valid gestational age code, we constructed a pregnancy window beginning 20 weeks after conception and ending on the delivery date. Steroid charges were considered valid exposures if:

- The charge matched the patient’s medical record identifier and hospital identifier
- The service date fell within the pregnancy window and on or before the delivery date

Steroid Identification

We include the following betamethasone formulations:

- Betamethasone sodium phosphate
- Betamethasone sodium phosphate/acetate combination products
- Celestone suspensions (1 mL and 5 mL vials)

We excluded topical formulations used for dermatological conditions, long-acting repository formulations, and preparations with non-standard dosing or administration routes inconsistent with antenatal corticosteroid protocols.

Exposure Timing and Classification

For each patient, we recorded all distinct steroid administration rates within the pregnancy window and calculated the number of exposures and days between each exposure and delivery. A patient was classified as exposed if at least one valid antenatal betamethasone charge was identified.

Defining a proxy variable for Level III NICU or higher neonatal designation of care

We constructed a proxy indicator for Level III NICU or higher neonatal intensive care unit designation based on the volume of extremely preterm births. First, we identified neonatal encounters for infants aged 0 years or classified under the newborns and neonates diagnostic category. Using *International Classification of Diseases, Tenth Revision, Clinical Modification* (ICD-10-CM) codes for disorders related to short gestation and low birth weight, we identified neonates with gestational age less than 32 weeks, including codes for extreme immaturity (generally 28 weeks or less) and preterm newborns with specific week designations (29 to 31 weeks). Second, we linked neonatal encounters to maternal deliveries at the same hospital within a narrow time window. The linkage criteria required matching hospital identifiers and infant admission dates within 1 day before or after the maternal delivery date. We retained only the first match per infant when multiple potential maternal deliveries were present. A hospital was designated as having Level III or higher neonatal intensive care unit capability each year if it had 10 or more neonates with gestational age less than 32 weeks linked to maternal deliveries. Hospitals with an average of 10 or more births per year were classified as Level III or higher facilities.

Categorization of Hospital-Level Steroid Use

We classified hospitals into quartiles based on their ALPS adoption rate during the post-ALPS period. This classification allowed us to examine heterogeneity in practice patterns and identify early versus late adopters of the ALPS trial recommendations. For each hospital, we calculated a weighted average adoption rate across the post-ALPS period using the following steps:

1. For each hospital and year in the post-ALPS period, we calculated the proportion of late preterm births that received ALPS
2. We compute a weighted mean of the annual adoption rates, with weights proportional to the number of late preterm births in each year.
3. The weighted mean adoption rates were rank ordered and separated into quartiles for comparison.

eFigure 1: Consolidated Standard of Reporting Trials Flow Diagram.

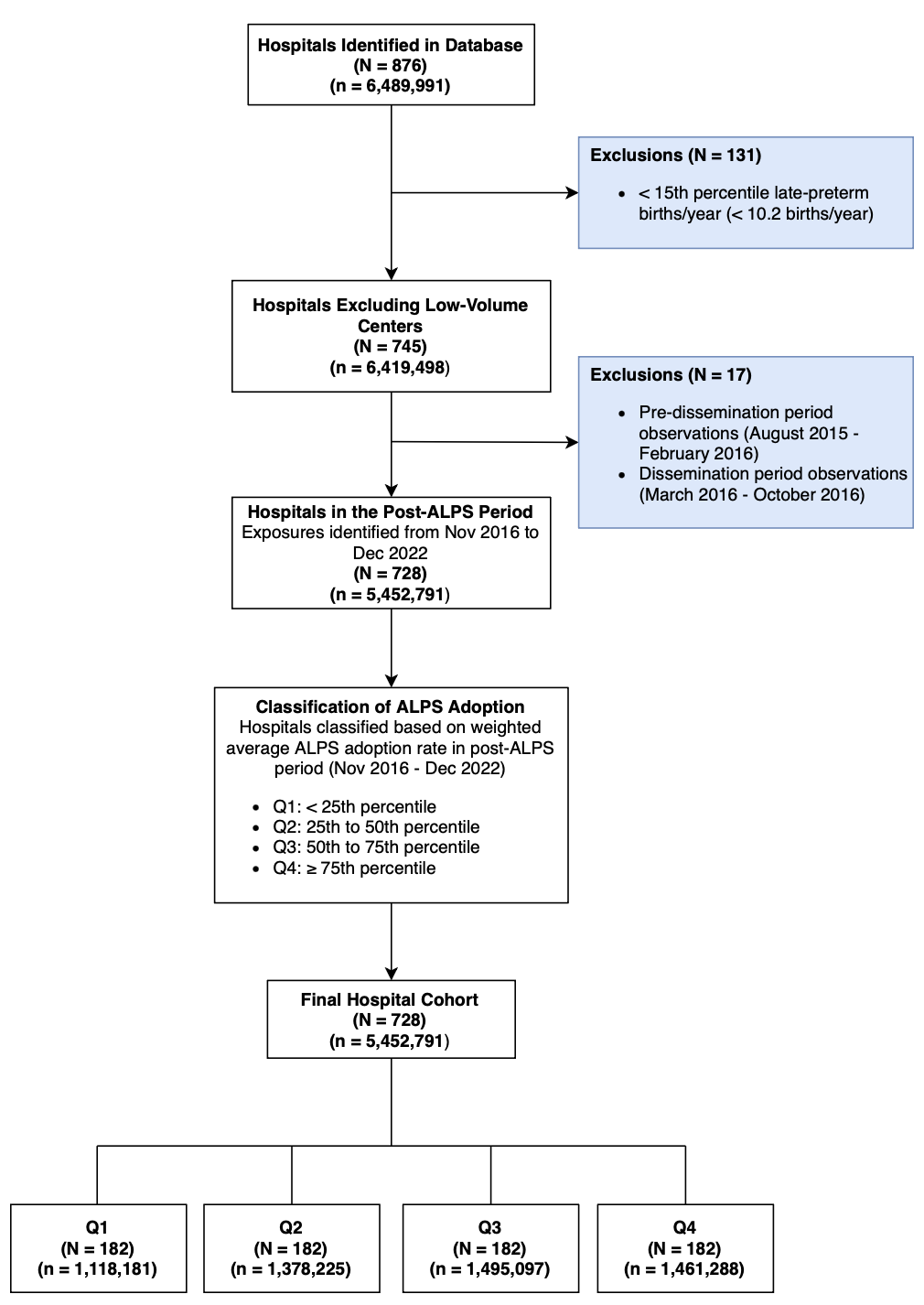

Hospitals were identified from the Premier Healthcare Database and assessed for inclusion after linking patient, billing, diagnosis, and procedure records. Exclusions included the post-trial dissemination period, low-volume hospitals, and no late-preterm steroid exposure in the post-ALPS period. The final cohort was classified into quartiles based on ALPS adoption rates among singleton deliveries.

eTable 1: ICD-10 and CPT Code List

| Category | Code Type | Code |
| --- | --- | --- |
| Maternal Delivery | ICD-10-PCS | 10E0, 10D0  (Obstetrics: Delivery and Extraction) |
| Vaginal Delivery | CPT | 59409, 59410, 59612, 59614 |
| Cesarean Delivery | CPT | 59514, 59515, 59620, 59622 |
| Maternal Gestational Age | ICD-10-CM | Z3A.XX (Weeks of gestation) |
| Early Preterm (24-33 weeks) | ICD-10-CM | Z3A.24 - Z3A.33 |
| Late Preterm (34-36 weeks) | ICD-10-CM | Z3A.34 - Z3A.36 |
| Term (37-42 weeks) | ICD-10-CM | Z3A.37 - Z3A.42 |
| Neonatal Gestational Age  (< 32 weeks) | ICD-10-CM | P07.21 - P07.26,  P07.31 - P07.34 |
| Preterm Premature Rupture of Membranes (PPROM) within 24 hours | ICD-10-CM | O42.01 |
| Multiple Gestation | ICD-10-CM | O30.* |
| Maternal Chronic Hypertension | ICD-10-CM | O10.*, I10, I11, I12, I13, I15 |
| Gestational Hypertension/Pre-Eclampsia | ICD-10-CM | O13.*, O14.*, O15.* |
| Pre-existing Diabetes (Type I/II) | ICD-10-CM | E10.*, E11.*, O24.1, O24.2, O24.3, O24.8 |
| Gestational Diabetes | ICD-10-CM | O24.4 |
| Preterm Labor | ICD-10-CM | O60.* |
| History of Preterm Delivery | ICD-10-CM | Z87.51, O09.21 |

***** Indicates all subcodes within the specified ICD-10-CM category.

eTable 2: Hospital characteristics associated with highest adoption of late preterm antenatal steroids.

| Characteristic | Unadjusted OR (95% CI) | Adjusted OR (95% CI) |
| --- | --- | --- |
| Urban | 3.24 (1.93-5.58) | 2.05 (1.11-3.83) |
| Teaching Hospital | 2.01 (1.31-3.09) | 1.02 (0.59-1.77) |
| Births per Month | 1.23 (1.02-1.52) | 0.94 (0.70-1.23) |
| Northeast | 1.24 (0.64-2.41) | 0.99 (0.48-2.06) |
| South | 0.42 (0.24-0.72) | 0.37 (0.20-0.69) |
| West | 0.93 (0.49-1.79) | 0.99 (0.47-2.06) |
| Level III NICU or Higher | 2.16 (1.40-3.36) | 1.03 (0.49-2.16) |
| Prop. GA 34 weeks | 1.95 (1.56-2.47) | 2.21 (1.58-3.14) |
| Prop. PPROM/Preterm Labor | 0.97 (0.79-1.19) | 1.18 (0.92-1.53) |
| Prop. Multiple Gestation | 1.29 (1.05-1.59) | 0.88 (0.68-1.16) |
| Prop. Type I/II Diabetes | 1.34 (1.09-1.66) | 1.01 (0.78-1.31) |
| Prop. Preterm Delivery History | 0.92 (0.65-1.11) | 0.87 (0.51-1.10) |

Adjusted odds-ratios were calculated using a logistic regression model comparing hospitals in the highest adopter quartile (Q4) to those in the lowest adopter quartile (Q1). The model adjusted for all variables shown. Reference groups for categorical variables include: Midwest region, non-teaching status, and rural location. Births per month and proportions for gestation age 34 weeks, PPROM/preterm labor, multiple gestation, diabetes, and preterm delivery history were z-score normalized.

eTable 3: Hospital characteristics associated with the highest adoption of late preterm antenatal steroids excluding maternal transfers.

| Characteristic | Unadjusted OR (95% CI) | Adjusted OR (95% CI) |
| --- | --- | --- |
| Urban | 3.09 (1.85-5.28) | 2.01 (1.10-3.75) |
| Teaching Hospital | 1.96 (1.28-3.01) | 1.03 (0.59-1.77) |
| Births per Month | 1.23 (1.02-1.52) | 0.94 (0.71-1.23) |
| Northeast | 1.16 (0.60-2.25) | 0.91 (0.44-1.89) |
| South | 0.43 (0.25-0.74) | 0.38 (0.20-0.70) |
| West | 0.93 (0.49-1.79) | 0.99 (0.47-2.07) |
| Level III NICU or Higher | 2.16 (1.40-3.36) | 1.07 (0.52-2.23) |
| Prop. GA 34 weeks | 1.92 (1.54-2.42) | 2.10 (1.53-2.94) |
| Prop. PPROM/Preterm Labor | 0.96 (0.78-1.18) | 1.17 (0.91-1.52) |
| Prop. Multiple Gestation | 1.29 (1.05-1.59) | 0.89 (0.67-1.16) |
| Prop. Type I/II Diabetes | 1.36 (1.10-1.69) | 1.04 (0.81-1.35) |
| Prop. Preterm Delivery History | 0.93 (0.65-1.11) | 0.88 (0.52-1.11) |

Adjusted odds ratios were calculated using a logistic regression model comparing hospitals in the highest adopter quartile (Q4) to those in the lowest adopter quartile (Q1), excluding patients who transferred facilities. The model adjusted for all variables shown. Reference groups for categorical variables include: Midwest region, non-teaching status, and rural location. Births per month and proportions for gestation age 34 weeks, PPROM/preterm labor, multiple gestation, diabetes, and preterm delivery history were z-score normalized.

eTable 4: Characteristics of the excluded low-volume hospitals and cohort included in primary analysis

| Characteristics | No. (%)  Overall Population  (N = 876) | No. (%)  Excluded  (N = 131) | No. (%)  Included  (N = 745) | P-value |
| --- | --- | --- | --- | --- |
| Urbanicity (%) | |  |  | <0.001 |
| Rural | 218 (24.9) | 69 (52.7) | 149 (20.0) |  |
| Urban | 658 (75.1) | 62 (47.3) | 596 (80.0) |  |
| Teaching Hospital (%) | |  |  |  |
| No | 559 (63.8) | 101 (77.1) | 458 (61.5) |  |
| Yes | 317 (36.2) | 30 (22.9) | 287 (38.5) |  |
| Provider Region (%) | |  |  |  |
| Midwest | 238 (27.2) | 49 (37.4) | 189 (25.4) |  |
| Northeast | 128 (14.6) | 15 (11.5) | 113 (15.2) |  |
| South | 344 (39.3) | 34 (26.0) | 310 (41.6) |  |
| West | 166 (18.9) | 33 (25.2) | 133 (17.9) |  |
| Late Preterm Births per Month (mean (SD)) | 8.45 (9.58) | 0.56 (0.43) | 8.89 (8.74) | <0.001 |
| Total Births per Month (mean (SD)) | 117.16 (116.70) | 14.12 (11.56) | 135.27 (117.45) | <0.001 |
| Prop. Late Preterm at 34 Weeks (%) (mean (SD)) | 13.76 (7.55) | 8.29 (12.24) | 14.59 (6.17) | <0.001 |
| Prop. Late Preterm with PPROM or Preterm Labor (%) (mean (SD)) | 49.46 (12.77) | 52.60 (20.32) | 48.99 (11.15) | <0.01 |
| Prop. Late Preterm with Multiple Gestation (%) (mean (SD)) | 7.99 (3.90) | 4.74 (5.97) | 8.48 (3.22) | <0.001 |
| Prop. Late Preterm with Type I or II Diabetes (%) (mean (SD)) | 2.95 (4.28) | 2.43 (10.70) | 3.02 (2.00) | 0.17 |
| Prop. Late Preterm with History of Preterm Delivery (%) (mean (SD)) | 1.23 (2.85) | 1.57 (3.66) | 1.18 (2.70) | 0.17 |

Differences in the continuous variables were assessed using analysis of variance and chi-squared tests for the categorical variables. The p-values are for the comparison between the first and fourth quartiles of steroid adoption rates.

eFigure 2: Trends in steroid exposure in late preterm births by adoption level including low-volume hospitals

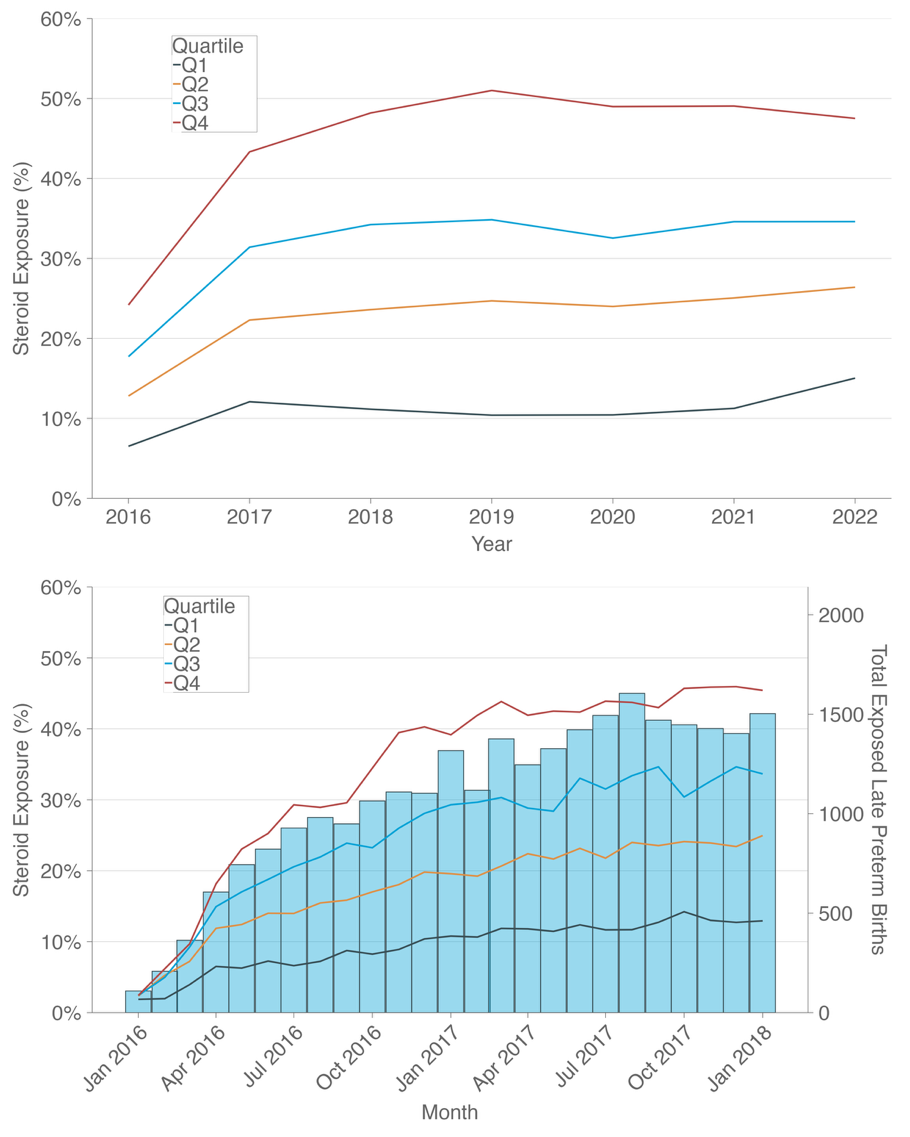

Yearly time series plot for steroid exposure among late preterm births and monthly time series plot for steroid exposure among late preterm births in the first two years stratified by quartile show a rapid and sustained adoption rate for Q4 hospitals and a modest increase in uptake for Q1 hospitals following the ALPS trial publication.

eTable 5: Hospital characteristics associated with highest adoption of late preterm antenatal steroids including low-volume hospitals

| Characteristic | Unadjusted OR (95% CI) | Adjusted OR (95% CI) |
| --- | --- | --- |
| Urban | 3.15 (1.98-5.13) | 2.20 (1.26-3.91) |
| Teaching Hospital | 2.10 (1.41-3.15) | 1.09 (0.63-1.87) |
| Births per Month | 1.42 (1.17-1.76) | 0.97 (0.74-1.26) |
| Northeast | 1.40 (0.77-2.57) | 0.85 (0.42-1.73) |
| South | 0.57 (0.35-0.92) | 0.41 (0.23-0.73) |
| West | 0.86 (0.49-1.53) | 0.75 (0.38-1.46) |
| Level III NICU or Higher | 2.28 (1.49-3.53) | 0.89 (0.44-1.80) |
| Prop. GA 34 weeks | 1.76 (1.42-2.21) | 2.28 (1.62-3.27) |
| Prop. PPROM/Preterm Labor | 0.85 (0.71-1.02) | 1.05 (0.82-1.36) |
| Prop. Multiple Gestation | 1.46 (1.20-1.79) | 1.04 (0.79-1.35) |
| Prop. Type I/II Diabetes | 1.01 (0.87-1.21) | 0.97 (0.64-1.32) |
| Prop. Preterm Delivery History | 0.87 (0.63-1.05) | 0.82 (0.52-1.08) |

Adjusted odds-ratios were calculated using a logistic regression model comparing hospitals in the highest adopter quartile (Q4) to those in the lowest adopter quartile (Q1). The bottom 15^th^ percentile hospitals that were excluded in the primary analysis were included in this analysis. The model adjusted for all variables shown. Reference groups for categorical variables include: Midwest region, non-teaching status, and rural location. Births per month and proportions for gestation age 34 weeks, PPROM/preterm labor, multiple gestation, diabetes, and preterm delivery history were z-score normalized.
